# Constellation illuminates rare disease genetics

**DOI:** 10.1101/2025.10.15.25337675

**Authors:** Siyuan Cheng, Qing Zhang, Xinchang Zheng, Shalini Jhangiani, Jacqueline C Weir, Jesse R Farek, Heer H Mehta, Ziad M Khan, Yi Han, Huyen H Dinh, Kerstin P Blankenburg, Jennifer E Posey, Richard A Gibbs, Donna M Muzny, Claudia M.B. Carvalho, Mitchell A Bekritsky, Ali Crawford, Daniel Calame, James Han, Fritz J Sedlazeck

**Affiliations:** Human Genome Sequencing Center, Baylor College of Medicine, Houston, TX, USA; Illumina, Inc., San Diego, CA, USA; Department of Molecular and Human Genetics, Baylor College of Medicine, Houston, TX, USA; Pacific Northwest Research Institute (PNRI), Seattle, WA 98122, USA; Section of Pediatric Neurology and Developmental Neurosciences, Department of Pediatrics, Baylor College of Medicine, Houston, TX, USA; Department of Computer Science, Rice University, 6100 Main Street, Houston, TX, USA

## Abstract

Despite significant advances in genomic sequencing, the resolution of many rare disease cases is still hindered by variant detection limitations. Short reads struggle in homologous regions, and long reads remain costly and difficult to scale. Here, we present the first systematic evaluation of Illumina’s Constellation sequencing technology for rare disease research. By fragmenting long DNA molecules directly on the flow cell surface, Constellation unlocks proximity information that enables long-range phasing and structural variant detection. Across 21 families, Constellation independently identified all known causative variants and resolved previously unsolved trios. It reliably resolved complex structural and copy number variants (e.g. impacting *MECP2*) and recovered haplotype phasing information across key disease impacting variants, all from low DNA input using existing Illumina infrastructure. These findings establish Constellation as a scalable, cost-efficient advance, closing critical diagnostic gaps and broadening access to long-range variant analysis in rare disease genomics.

## Introduction

Over the past two decades, DNA sequencing has transformed our ability to identify disease-causing variants in rare and undiagnosed genetic conditions, while simultaneously mapping genetic diversity and phenotypic drivers across populations^1–3^. Multiple technical innovations have enabled this transformation, including short-read DNA sequencing (∼32-150bp) and ultra-long read DNA sequencing (with up to 4Mbp), which have improved analysis, comprehensiveness of variant detection, scalability, and affordability^1^. Illumina short-read sequencing remains the workhorse of genomics, with established pipelines and cost-efficient approaches for small– and large-scale sequencing^1^. Despite this progress, the diagnostic yield for genetic disease remains below 40%^2^, highlighting a considerable opportunity for further technical improvement.

This is in part due to the fact that the technical challenges for improved genomic analyses extend beyond detection of single nucleotide variants (SNVs) and small insertions or deletions. The field has since expanded to capture more complex variant classes, including structural variants (SVs), copy number variants (CNVs), and tandem repeats (TRs)^4–6^. Despite these advances, no single technology has yet achieved comprehensive detection across all variant types while maintaining cost efficiency and scalability. Third-generation long-read platforms, such as Pacific Biosciences (PacBio) and Oxford Nanopore Technologies (ONT), have substantially improved the resolution of complex alleles, repetitive regions, and methylation signatures while enabling haplotype phasing to interpret compound genetic effects^7,8^. However, their high costs and DNA input requirements have limited widespread adoption at the scale of short-read exome or genome sequencing^9^. In parallel, newer short-read platforms such as Aviti^10^ and Ultima Genomics^11^ aim to increase sequencing accuracy or lower per-genome costs, respectively. Yet, none of the current approaches effectively combine affordability, comprehensive variant detection, scalability, and minimal DNA input, which highlights the continued need for innovative sequencing technologies that deliver long-range insights within an accessible, high-throughput framework.

Most recently, Illumina announced Constellation mapped read technology (For Research Use Only), which is a novel approach that enables efficient proximity read sequencing without extensive laboratory preparation, PCR, or DNA modification. Previous linked read approaches, such as 10x Genomics or TellSeq^12^, also showed great potential to enable more comprehensive genome sequencing, but required arduous DNA library preparation, PCR, or DNA modification. Constellation differs as it loads long, unfragmented DNA directly onto the flow cell for fragmentation, and records the location of each standard Illumina paired-end read during sequencing. This location information is used during analysis to identify paired-end reads that are likely to belong to the same DNA molecule. Constellation leverages the physical colocation of reads on the flow cell to create spatially proximal sets of reads derived from contiguous genomic DNA molecules spanning tens to hundreds of kilobases (**Figure 1A**). This information enhances the insight delivered by short read genomes, including enabling phasing and improving mapping in difficult-to-map regions and variant calling. Constellation requires only nanogram-scale DNA input and maintains high data yield even from suboptimal samples, offering exceptional flexibility in DNA quality while preserving coverage (**Figure 1B, Supplementary Figure 1**).

**Figure 1.**
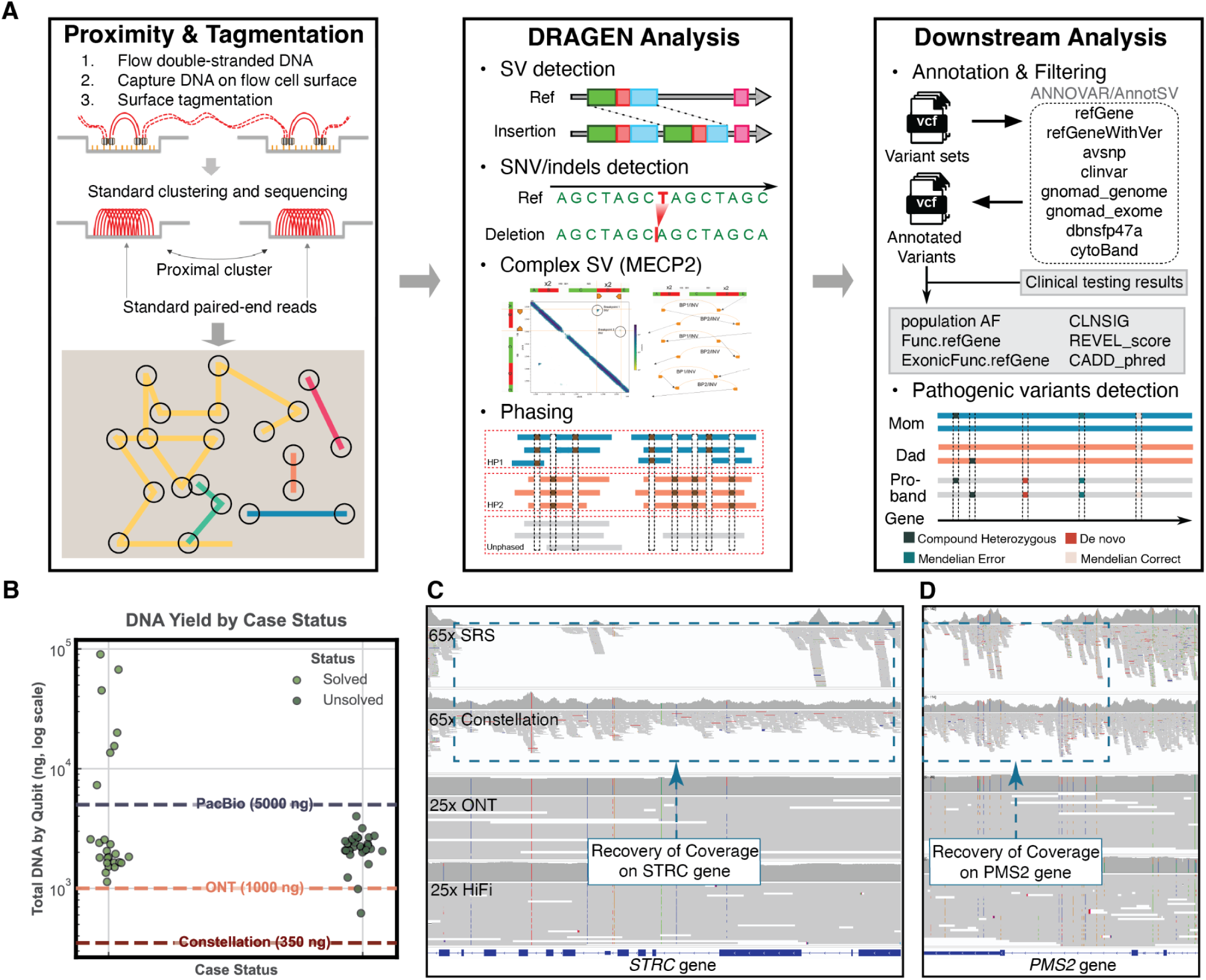
Experimental design of the Constellation benchmark analysis. **(A)** Schematic overview of experimental workflow. Constellation sequencing enables long-range phasing and variant calling through on-flow-cell tagmentation and proximity information, with downstream annotation performed (**Methods**). **(B)** DNA amount available for each sample in solved and unsolved trios. Different colors represent solved (light green) and unsolved trios (dark green). As illustrated, only Constellation was able to sequence most of these samples without completely depleting the available DNA. Input requirements for Constellation at 350ng, ONT at ∼1ug^15^ and PacBio at ∼5ug^16^. **(C)** Improved mapping in high homology regions. Recovery of coverage in the STRC genes. **(D)** Recovery of coverage in the PMS2 genes.

In this paper, we assess the potential of Constellation technology for variant calling in rare disease research. We first extensively benchmarked this new technology against standard short– and long-read (ONT and PacBio) sequencing using the Ashkenazi Jewish GIAB reference trio (HG002, HG003 and HG004)^13^. We observed advantages of Constellation when compared to long-reads for SNV and indel calling, and short-reads for SV calling. Motivated by these results, we next assessed Constellation’s ability to identify causative variants in rare disease cases from the Genomics Research to Elucidate the Genetics of Rare Diseases (GREGoR) Consortium^13,14^. First, we identified cases with known compound heterozygous variants to assess Constellations phasing capabilities in tissue samples from study participants. Secondly, we evaluated this new technology’s ability to detect highly complex variants, such as multibreakpoint structural rearrangements observed in individuals with *MECP2* duplication syndrome (MIM#300260). Constellation was able to resolve these challenging cases, though final confirmation required manual review. Our approach (**Figure 1A**) was able to independently solve all previously solved cases. Finally, we evaluated Constellation in unresolved cases that had previously undergone clinical and research sequencing. These are clinically challenging cases with variants that escaped established research and even some clinical sequencing. For this manuscript, we highlight two such cases that we were able to solve using Constellation.

## Results

### Comprehensive benchmarking on the Constellation sequencing

The Constellation workflow is summarized in **Figure 1A**. Unfragmented DNA is loaded directly into the library strip tube and fragmented on the flow cell by the sequencing instrument after run initiation. **Supplementary Section 1** discusses the impact of DNA quality. The physical location of each fragment is recorded from the flow cell (left panel) and used to generate proximity information. After sequencing, the data were analyzed on the Illumina Connected Analysis (ICA) platform, and variants were called and phased with DRAGEN for SNVs and SVs. Proximity information noticeably improved mapping and variant calling in complex regions, which is achieved by uniquely mapping ambiguously aligned reads to the correct location using uniquely mapped reads within the same proximity group (**Figure 1A**, middle panel). We implemented a custom workflow to rank and annotate variants to foster rare disease discovery (**Figure 1A**, right panel; **Methods**). We assessed the performance of Illumina Constellation technology for small variant calling accuracy on four replicates of HG002, and for phasing and Mendelian consistency on a combined analysis of two HG003 (Father) and HG004 (Mother) replicates.

We first examined Constellation’s usage of proximity signals from adjacent, uniquely mapped clusters to improve read mapping. We observed more confident mapping and comprehensive coverage of the genome, including difficult-to-map, medically relevant genes such as *STRC* and *PMS2*, compared to standard short-read DNA sequencing (**Figure 1C, D**). The sequence identity for both *STRC* and *PMS2* with their pseudogenes, *STRCP1* and *PMS2CL*, exceeds 99%, making mapping challenging with standard short-read DNA sequencing. Genome-wide SNV and indel detection performance showed a high F-score and high consistency across the four replicates (F1 score SDs = 0.000096 on the GIAB T2T-Q100 v1.1 benchmark set with genotype match requirement) when aligned against the GRCh38 reference genome (**Figure 2A**). Against the GIAB v4.2.1 benchmark callset (GRCh38-based)^17^, we recovered on average 3,888,264 ± 65 true-positive variants (SNVs+indels), with a mean F1 of 0.9995 (**Supplementary Table S1**). On the GIAB T2T-Q100 v1.1 benchmark set^18^, Constellation achieved a mean true positives 4,489,991 ± 473 (SNVs+indels), with F1 of 0.9952 (**Supplementary Table S1**). Lastly, in the challenging CMRG v1.0 benchmark^19^, we observed high performance despite the repetitive content, yielding 21,026± 8 true-positive SNVs+indels, with F1 of 0.9929 when requiring GT match (**Supplementary Table S1**). The comparison between the two GIAB whole genome benchmarks highlights critical differences across the benchmark sets: while average F1 scores of GIAB T2T-Q100 were 0.0043 lower than GIAB v4.2.1, it identified massively more true positives (average 601,727 for SNV+indels). In addition, a small subset of variants that were classified as false positives in the GIAB v4.2.1 were reclassified as true positives in the GIAB T2T-Q100 set, averaging 292 ± 11 events for SNVs+indels across the four replicates. Overall, Constellation shows high performance across all three GIAB benchmarks.

**Figure 2.**
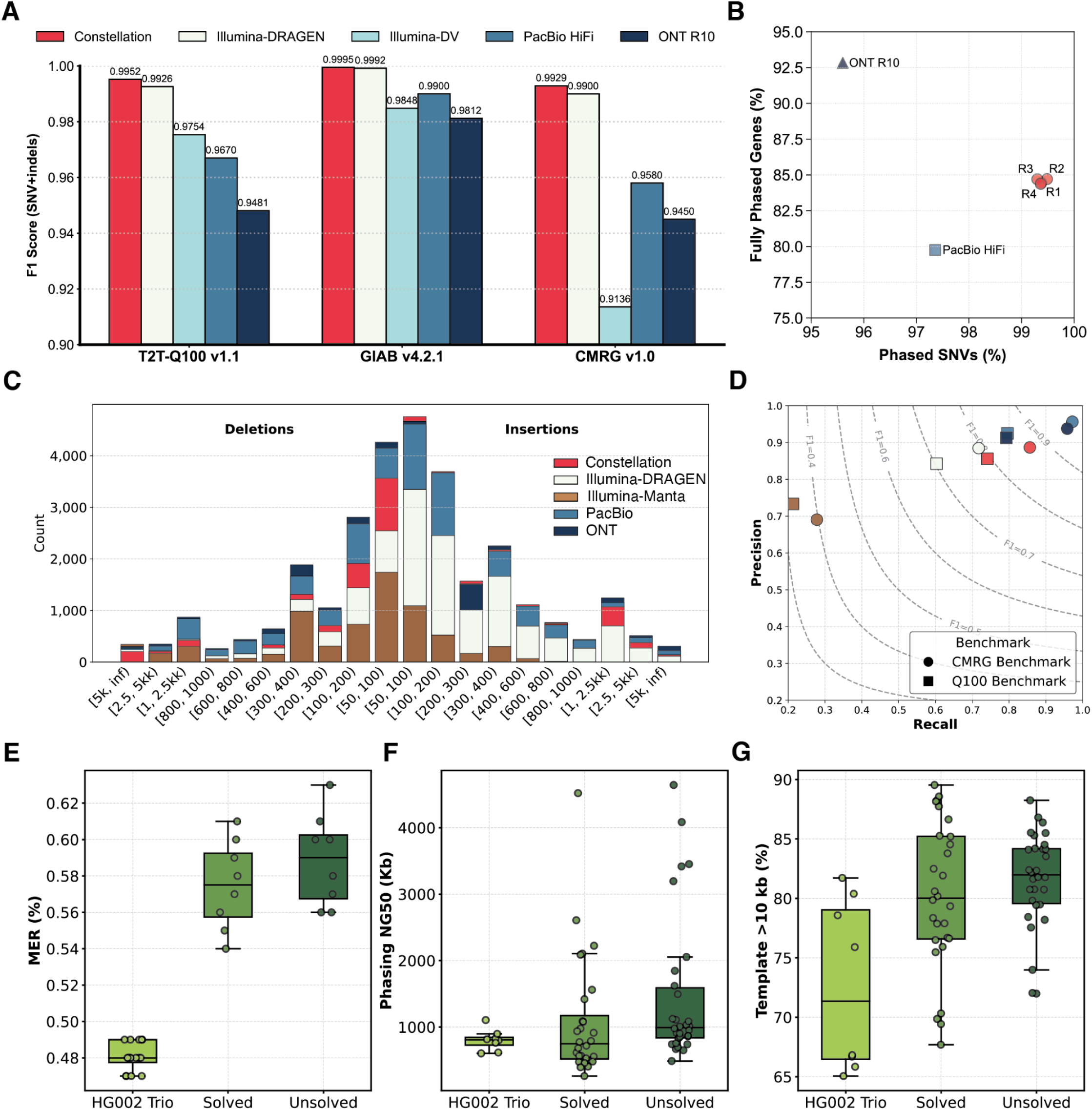
Comprehensive benchmarking of Constellation relative to short– and long-read technologies on the HG002 sample. **(A)** Single-nucleotide variant (SNV) and indel accuracy. F1-scores across three independent benchmark sets (GIAB T2T-Q100 v1.1, GIAB v4.2.1, and CMRG v1.0). Illumina-DV: Illumina short-read with DeepVariant; PacBio HiFi and ONT R10: LRS data with Sniffles2. **(B)** Haplotype phasing performance. Scatterplot of phased SNV fraction versus percentage of fully phased genes. **(C)** Size spectrum of indel detection. Distribution of insertion and deletion calls of 5 sequencing and calling methods across binned size categories. **(D)** Precision-recall-f1 curve. Precision-recall plots across CMRG v1.0 and Q100 benchmarks, overlaid with F1 contours. **(E)** Mendelian Error Rate (MER) across HG002 trios (light green, n=16), solved (green, n=8), and unsolved trios (dark green, n=8). **(F)** Phasing NG50 length across HG002/3/4 samples (n=8), solved trios (n=28), and unsolved trios (n=30). **(G)** Percentage of DNA templates that are longer than 10 Kb across HG002/3/4 samples (n=8), solved trios (n=28), and unsolved trios (n=30).

We further benchmarked Constellation technology against 30x standard Illumina short-read DNA sequencing technology (SRS) from precisionFDA, 25x PacBio HiFi, and 25x ONT R10 (both data come from GIAB) long-read sequencing technology (LRS) for SNV/indels detection. On SRS, Constellation consistently outperformed DeepVariant^20^ SNV calling with standard Illumina short-read sequencing in SNV+indels calling with F1=0.985 (GIAB v4.2.1), 0.975 (GIAB T2T-Q100 v1.1), and 0.917 (CMRG v1.0), respectively (**Figure 2A**). We next investigated whether the graph genome from DRAGEN^21^ for SRS can directly improve overall performance (**Supplementary Table S1**). Constellation outperformed DRAGEN with SRS for all benchmarks with SRS performance of F1=0.9992 (GIAB v4.2.1), 0.9926 (GIAB T2T-Q100 v1.1), and 0.9900 (CMRG v1.0), respectively. Long-read data using Clair3^22^ exhibited large degradation in SNV+indels calling: PacBio HiFi achieved F1 scores of 0.9896 (GIAB v4.2.1), 0.9674 (GIAB T2T-Q100 v1.1), and 0.9592 (CMRG v1.0), whereas ONT R10 trailed a bit with F1 scores of 0.9812, 0.9481, and 0.9450 on the same benchmark sets, respectively. **Supplementary Table S1** shows all the details on comparisons for SNVs and indels.

We next assessed Constellation technology’s phasing ability, which is enabled by the proximity maps of reads along the flow cell. Here in four HG002 replicates, we observed a mean phasing NG50 of 800.4 Kb (SD=23.6 Kb) and the longest phasing block of 8.7 Mb (**Figure 2F**) with consistently assigned a high fraction of heterozygous SNVs, 99.38% ± 0.08% (Mean ± SDs), to a haplotype (**Figure 2B**). It enabled an average of 84.56% genes to be fully phased by a single phasing block (**Figure 2B**). We also validated phasing accuracy against the reference dataset, with an average switch error rate of 0.062% ± 0.0017% across all blocks and 0.033% ± 0.006% within the longest blocks (**Methods**). Compared with long-read platforms on HG002, Constellation was better than PacBio Revio’s single run result in contiguity (longest phasing block: 4.08 Mb, NG50: 441 Kb), phased SNV fraction (97.36%), and gene-spanning (79.76%). In contrast, ONT R10 provided greater contiguity (longest phasing block 9.33 Mb, NG50: 1.78 Mb), better gene coverage (92.83%), but slightly lower SNV fraction (95.60%) (**Figure 2B**). Besides, using HG003 and HG004 replicates, we benchmarked the Mendelian error rate (MER), a key metric for rare disease studies, and observed a remarkably low mean MER of 0.48% (SD = 0.007%) (**Figure 2E**).

Next, we assessed the performance on SV calling, which has been a consistent challenge for short-read DNA sequencing^23^. Constellation technology produced a final set of an average of 25,071 SVs (16,097 insertions and 8,975 deletions) across 4 replicates (**Figure 2C**). For the GIAB T2T-Q100 v1.1 benchmark, Constellation achieved a mean F1 score of 0.796 ± 0.002 (**Figure 2D**). Notably, when benchmarked against the CMRG v1.0 SV set, we observed a mean F1 score of 0.865 ± 0.01 (**Figure 2D**). Next, we compared this performance with the other technologies. Using the GIAB T2T Q100 v1.1 benchmark, for SRS, Manta^24^ achieved a mean F1 score of 0.330. This was improved by DRAGEN SV caller (F1=0.7029) (**Figure 2D**). Constellation improved upon these significantly (F1=0.796). As expected, LRS using Sniffles^5^ performed well with PacBio (F1=0.855) and ONT (F1=0.849) (**Figure 2D**). This performance hierarchy was consistent when using the CMRG v1.0 benchmark, where Constellation’s mean F1-score of 0.865 was lower than LRS technologies (PacBio: 0.964; ONT: 0.948) and again demonstrated a profound accuracy advantage over both DRAGEN (0.793) and Manta (0.396) (**Figure 2D**).

Thus, we observe that Constellation shows a high performance across all variant types and is able to phase these variants accurately. Based on our benchmark, it outperforms LRS and standard SRS in SNV and SV, respectively, and thus highlights its position as a cost-efficient alternative that requires much lower DNA inputs compared to LRS.

### Constellation recovers known pathogenic variants in rare disease trios

We next applied Constellation to a cohort of eight trios and one proband from the GREGoR project with characterized pathogenic variants to evaluate its performance on rare disease data. Notably, the limited amount of genomic DNA available from these samples highlights the potential of Constellation in rare disease genomics, where LRS requires a much higher DNA input amount (**Figure 1B**). In these trios, only 7/25 samples had enough DNA to be sequenced on PacBio. The DNA amounts were sufficient for WGS on ONT at the risk of depleting all the DNA, which leaves little for confirmation or future experiments. Across the 25 sequenced samples (eight trios and 1 proband only), we obtained a mean coverage of 66.4x (SD=3.1). On average, we detected 5,226,890 SNVs/indels (**Figure 3A**) and 25,026 SVs (SD=1,065) per sample (**Figure 3B**). Phasing metrics demonstrated robust haplotype reconstruction, with an average phasing NG50 of 1.08 Mb and 98.3% (SD=0.3%) of heterozygous SNVs phased (**Figure 1B, 3C**). Additionally, 81.7% (SD=7.0%) of protein-coding genes were fully phased (**Figure 3C**). The single sample variant calling leads to a low Mendelian error rate (MER) of 0.58% (SD=0.02%) (**Figure 2B**).

**Figure 3.**
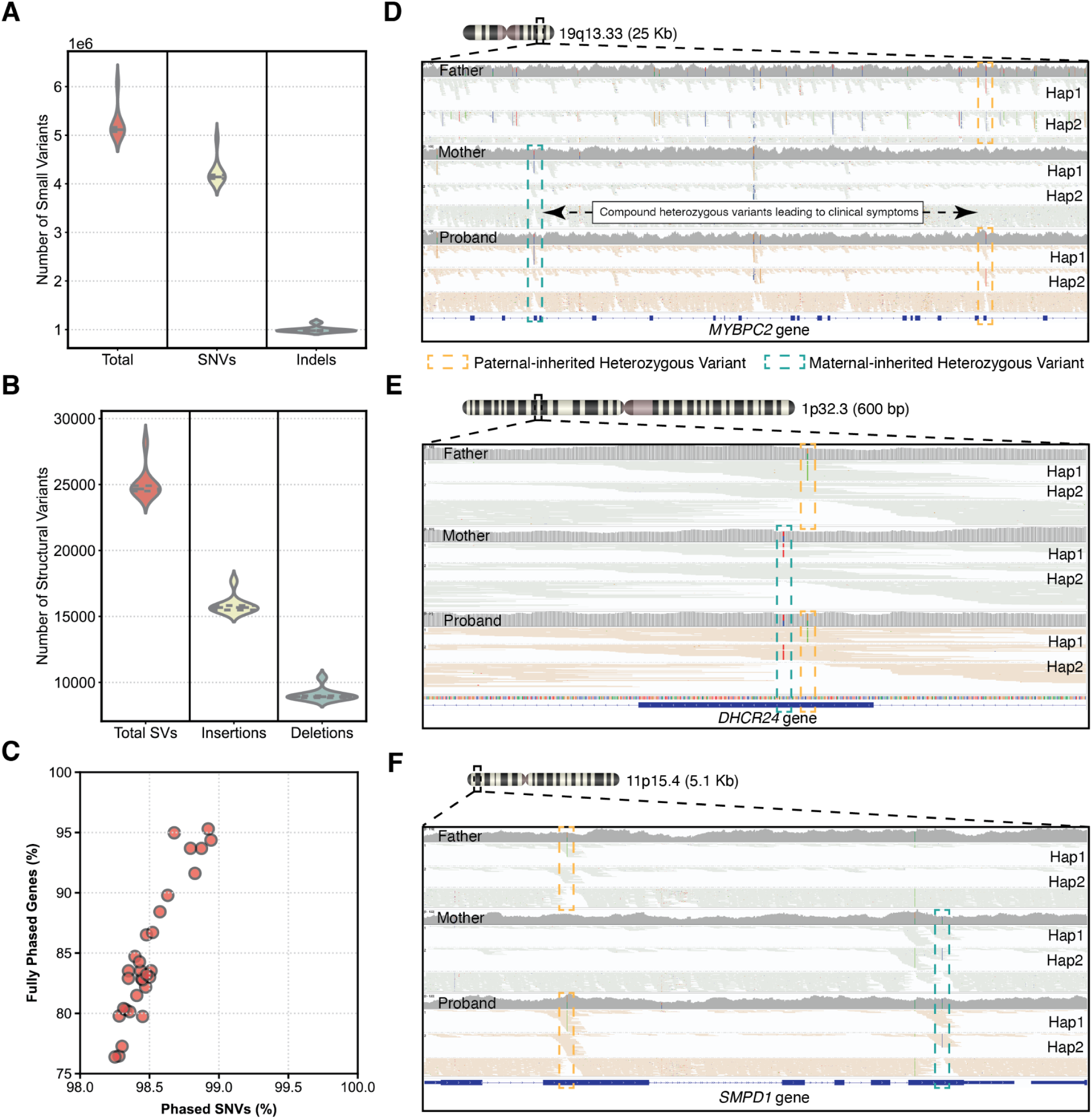
Application of Constellation on Solved Mendelian Trios. (**A, B**) Number of SNVs/indels and SVs called across 25 samples. **(C)** Haplotype phasing performance. Scatterplot of phased SNV fraction versus percentage of fully phased genes. **(D-F)** Causative compound heterozygous variants independently re-identified in family BH10120 **(D,** MYBPC2 gene), BH11320 **(E,** DHCR24 gene) and BH12598 **(F,** SMPD1 gene). The separation of reads on each sample representing different haplotypes (maternal and paternal). Maternal-inherited SNVs were labeled in green dashed lines, and paternal-inherited SNVs were labeled in orange dashed lines. Probands inherited one SNV from each parent and led to clinical disease.

First, we investigated 6 trios that were known to harbour 24 compound heterozygous causative variants consistent with their disease phenotypes (**Supplementary Table S3**). We applied our in-house annotation and filtering pipeline to identify likely pathogenic variants, both compound heterozygous variants and Mendelian error variants (**Methods**). Overall, we observed an average of 7.7 Mendelian-error (SD=8.9) and 54.8 compound heterozygous (SD=23.2) SNVs/indels events on exonic regions. Utilizing our analysis framework, we independently identified all 24 compound heterozygous pathogenic variants across the 6 trios (**Methods**). Furthermore, Constellation successfully phased 22 of the 24 heterozygous variant pairs, accurately resolving all parental origin (**Supplementary Table S3**). **Figure 3D** shows two heterozygous SNPs in the *MYBPC2* (NM_004533.4) gene, which were confirmed to be the causative allele for BH10120 with an identified arthrogryposis multiplex congenita and tapering fingers. Constellation showed clearly that the first variant, c.3194C>T (p.Ala1065Val) at chr19:50462002, was paternally inherited, while the second variant, c.920T>C (p.Val307Ala) at chr19:50443511, was maternally inherited. **Figure 3F** shows two heterozygous variants in the *SMPD1* (NM_000543.5) gene, which were also confirmed to be the causative variant for BH12598, given its phenotype (neurodegenerative disorder and developmental regression). The first variant, c.502G>A (p.Gly168Arg) at chr11:6391567, was inherited from the father and is documented as a pathogenic allele associated with type A Niemann-Pick disease and related sphingomyelin/cholesterol lipidosis phenotypes, as reported in ClinVar (982662, VCV000982662.10)^25^. The second variant, 11:6394441:A>C, also falls within an exon of *SMPD1*, but has not been reported in ClinVar. The same compound heterozygous variant pattern was verified in the BH11320 family (**Figure 3E**), concordant with a clinical phenotype of severe developmental delay, intellectual disability, hyperreflexia, and spasticity. These results demonstrate Constellation’s ability to phase pathogenic heterozygous variants, enabling accurate resolution of compound heterozygous inheritance patterns.

We next explored the performance of Constellation on SVs and complex repeat regions. These are part of more complex structures that are considered unsolvable with SRS and even challenging for LRS^5,23^. Nevertheless, a multi-technology focused effort could resolve several mechanisms underlying complex SV impacting *MECP2*^26,27^. From this effort, we sequenced three proband samples with increasing complexity using Constellation (BH14233_1, BH13947_1, and BH15700_1). The challenge lies in the location of the *MECP2* gene, which maps to Chromosome Xq28, flanked by multiple segmental duplications, which cause mapping and variant calling challenges^26^. We previously showed the performance of Constellation on SV calling over the genome and indeed it reported multiple CNV and SV per sample. However, to fully resolve this region of *MECP2* requires manual investigation due to its complexity. Constellation can leverage the physical colocation of reads on the flow cell to interrogate genome architecture, summarized in a colocation plot, which encodes the number of proximity contacts between reads from each pair of genomic bins (**Figure 4A**, left panel). Colocation plots were generated for each sample in the region of interest. **Figure 4A** shows the patterns of the colocation plots for simple SVs, including deletion, insertion, inversion and tandem duplication. In the colocation plot, forward junctions (same orientation as read orientation LR or RL) are indicated by density on the first quadrant (Q1) and the third quadrant (Q3). In contrast, reverse junctions (inverted orientations with read names LL or RR) are indicated by the second quadrant (Q2) and the fourth quadrant (Q4) (**Figure 4B**). For the model figures in Figure 4: copy numbers for each segment are labelled on the X/Y axis. Green indicates copy number 1, salmon color indicates copy number 2, and burgundy color indicates copy number 3.

**Figure 4.**
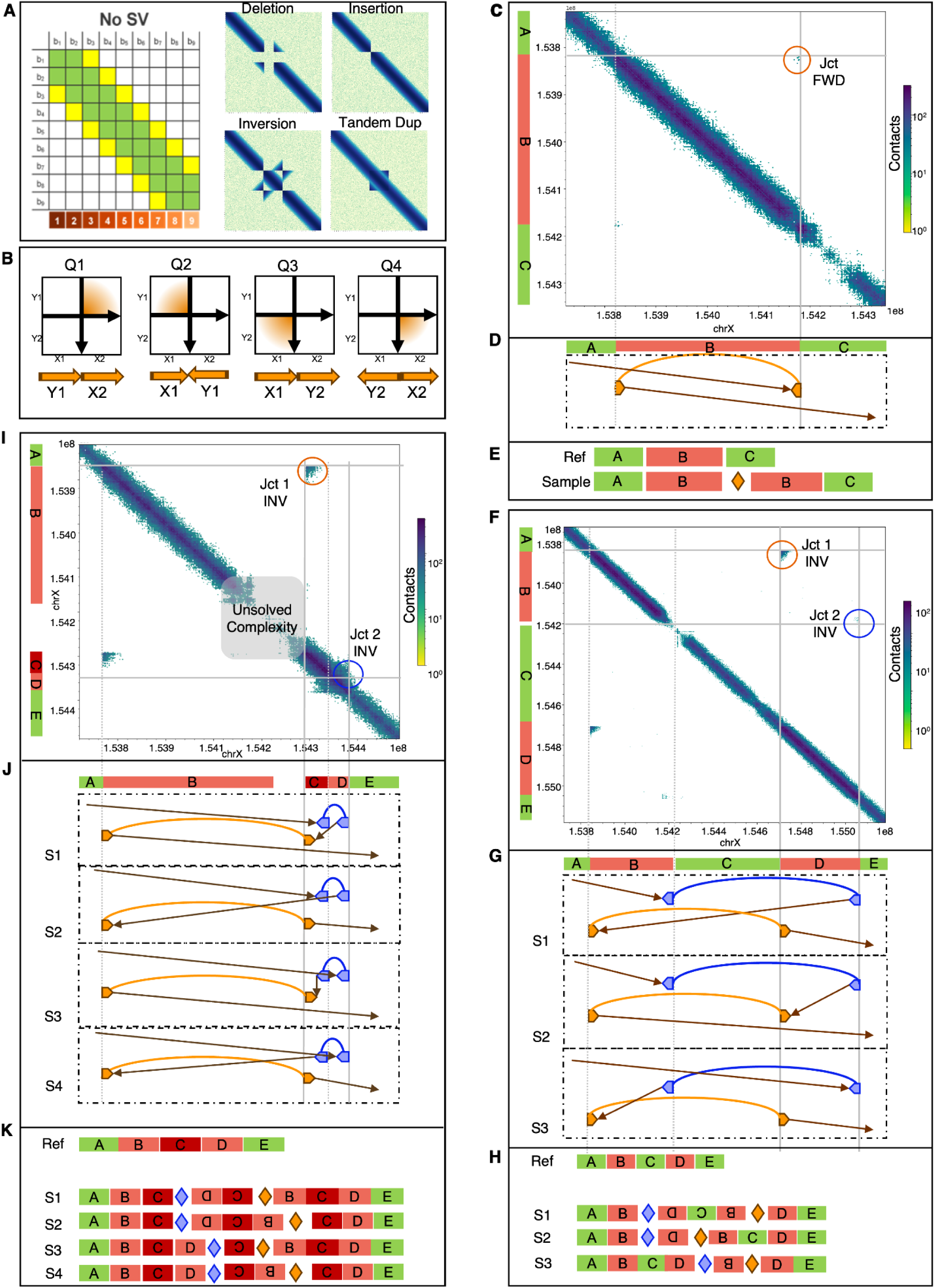
MECP2 Structures detected by Constellation in Mendelian disorders in probands. **(A)** Left: definition of colocation matrix. Right: Illustration of simple SVs on the colocation map. **(B)** Junction and orientation representations in the colocation matrix. Density on Q1/3 are “forward junctions” where the segments on either side follow the same directionality wrt. Reference; density on Q2/4 are “reverse junctions” where the segments on either side have opposite directionality wrt. reference **(C-E)** Colocation map, and proposed haplotype structure of the tandem duplication in proband of BH14233 family. **(C)** On the X/Y axis of the colocation map are the CNV tracks of each segment. Salmon color represents CN=2, green represents CN=1, and burgundy color represents CN=3. The same color scheme applies to colocation figures as F/I. **(D)** dotplot of the sample’s sequence aligning to the reference. If more than one scenario can fit with the CN profile and orientation of junctions in colocation plot, multiple dotplots and SV haplotypes will be shown and labelled as Scenario 1 (S1), Scenario 2 (S2), etc. A linear representation corresponding to each scenario is shown in **(E)**. **(F-H)** complex duplication-normal-duplication/inversion (DUP-NML-INV/DUP) structure in BH13947 proband. Three possible SV complex SV delineations were shown. **(I-K)** Colocation map of BH15700 proband with duplication-triplication-duplication/inversion (DUP-TRP/INV-DUP) structure. Four possible SV complex SV delineations were shown.

The simplest case in the BH14233 family. The CNV gain at segment B matches the increased contacts on the diagonal. There is also one off-diagonal junction (the orange circle) joining the end of B with the start of B with density in the third quadrant, suggesting that this is a forward junction. Several chimeric reads in that junction showed RL pair read orientation, further supporting that the junction is a forward junction. Combining the read signal and colocation signal, we confirmed that this is a tandem duplication of segment B (**Figure 4D**)^5^. We thus independently identified the causative variant for this proband, as it leads to an amplification of *MECP2,* a copy number sensitive gene^26^, a known molecular mechanism for the disease.

In the BH13947 proband, which is a more complex case, two CNV gains and two off-diagonal signals were called at regions B and D in the proband (**Figure 4 F-H**). Jct1 (**Figure 4F**, orange circle) joins the start of segment B with the start of segment B in inverse orientation, as density on Q4 and RR pair orientation in the SV breakends. The second junction is more subtle from colocation as it joins the end of segment D to the end of segment B, which is within a low-copy repeat (LCR). Read pair evidence suggested that the supporting read pairs are in LL orientation, making the second junction also an inverse junction. Given the orientation and location of the breakpoints and the copy number of each segment, there could be three complete SV haplotypes (**Figure 4G-H**, models S1-3) consistent with a duplication-normal-duplication (DUP-NML-DUP) structure and two overlapping inversions configuration, leading to the *MECP2* abnormality and disease phenotype.

The proband in the BH15700 family harbored another complex SV in the *MECP2* locus (**Figure 4I-K**). DRAGEN identified two CNV gains in this region. On the colocation map, two off-diagonal inverse junctions with segments of CNV gains were detected by Constellation. Junction 2 (Jct 2, blue circle) joins the end of region C with the end of region D. Junction 1 (Jct 1, orange circle) joins the beginning of region C to the beginning of region B inside the *L1CAM* gene, which is flanked by inverted segmental duplications (SDs). Although no chimeric reads can be identified in either junction in this case, the directionality and position of the junctions can still be deduced from the off-diagonal signals from the colocation plot arising from the proximity of reads from the same templates. This evidence gave rise to four possible haplotype configurations (**Figure 4J-K**, models: S1-S4) that agree with the duplication-triplication-duplication (DUP-TRP/INV-DUP) structure reported previously^5,26^.

Through the analysis of trios with previously characterized pathogenic variants, both compound heterozygous and complex events around *MECP2*, we demonstrate the ability of Constellation to resolve rare disease cases. For the *MECP2*, Constellation did not call all breakpoints automatically, but it remains remarkable that the SV breakpoints can be deduced from proximal contacts via colocation plot to allow delineation of complex SV events in difficult regions as *MECP2*.

### Constellation on rare disease families with unknown mutations

Given the encouraging results on Mendelian disease trios with known pathogenic variants, we next applied Constellation to a cohort of eight unsolved rare disease trios that were not solved with standard Illumina SRS. All undiagnosed samples went through comprehensive rare disease analysis, including exome and genome sequencing, while a subset also underwent research exome, genome, and RNA sequencing (**Supplementary Table S4**). Across the 30 sequenced samples, the cohort achieved a mean coverage of 54.0x (SD=5.8). Importantly, 2/30 samples can only be sequenced by Constellation, and none of the unsolved samples can be sequenced by PacBio (**Figure 1B**). On average, variant discovery yielded 5,220,580 SNVs/indels (SD=301,439) and 24,836 SVs (SD=1,034) per sample. Phasing metrics demonstrated strong haplotype resolution, with an average phasing NG50 of 1.47 Mb (SD=1.10 Mb) and 98.5% (SD=0.2%) of heterozygous SNVs successfully phased. Additionally, 84.8% (SD = 5.5%) of protein-coding genes were fully phased, highlighting the robustness of phasing performance across the dataset. Among eight complete trios, we observed a low average Mendelian error rate (MER) of 0.90% (SD=0.06%) with on average 5.8 violating variants and 52.4 compound heterozygous (SD=22.0) SNVs/indels (**Methods**). Based on the described phenotype and clinical information, we received a recommended gene list for each family from an expert clinician scientist and the Genomics England PanelApp^28^. We further filtered variants that overlapped with gene lists and verified with clinical information to identify undiagnosed or unreported pathogenic variants (**Methods**). As proof of concept, we want to highlight two cases below, especially.

In the BH13518 family, the proband has a metabolic myopathy with clinical untargeted plasma metabolomics showing acylcarnitine elevations consistent with multiple acyl-CoA dehydrogenase deficiency (MADD) or riboflavin metabolism disorders. His parents were related, and his sibling was similarly affected and died of disease complications in childhood. Clinical and research exome and genome sequencing failed to identify pathogenic variants in phenotypically relevant genes such as *ETFA, ETFB, ETFDH, FLAD1, SLC52A1, SLC52A2, or SLC52A3*. The shared parental ancestry and recurrence in a sibling strongly suggest an autosomal recessive inheritance pattern. Applying the in-house annotation and filtering pipeline on the sequencing result, one SNV, 4:158680528:A:G, was found to overlap with an exon of the *ETFDH* gene, but was predicted as a synonymous substitution (**Figure 5A**). However, the same variant was found in paired short-read RNA-seq data (srRNA-seq), indicating a clear alteration of splicing linked to the variant, thereby confirming its pathogenicity per ACMG criteria^29^ (**Figure 5B**). This was further supported by a high Splice AI^30^ score of 0.56. Review of prior research exome sequencing data demonstrated that the variant was present in the proband and parental BAM files but was absent from filtered CSV analysis files. The identification of pathogenic variants via novel technologies like Constellation, which could have been detected by exome sequencing data reanalysis, is a recurring theme in rare disease research. Therefore, we identified the treatable metabolic disorder Multiple Acyl-CoA Dehydrogenase Deficiency in this patient by combining Constellation and srRNA-seq data and identified a novel pathogenic *ETFDH* variant.

**Figure 5.**
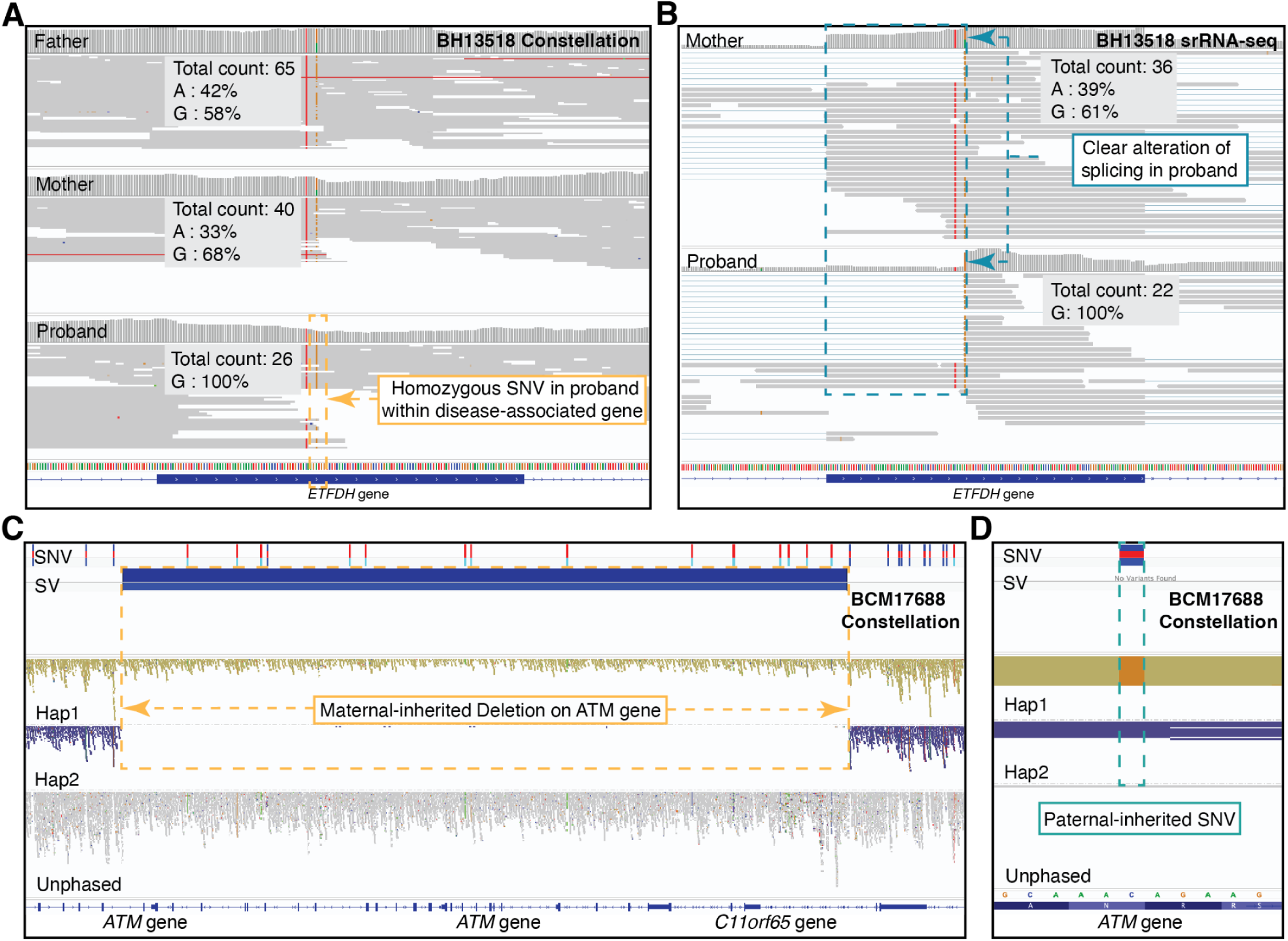
Application of Constellation on Unsolved Mendelian Trios. **(A)** Proband-specific homozygous SNV on the exon of the ETFDH gene, while parents are all heterozygous, in the BH13158 family. **(B)** The same variant found in Constellation showed a clear alteration of splicing in the proband from the paired srRNA-seq results in the BH13158 family. **(C)** Maternal-inherited deletion on the ATM gene in the BCM17688 family. **(D)** Paternally-inherited SNV on the exon of the ATM gene in the BCM17688 family. Colored dashed boxes highlight variants with their matching description.

In the BCM17688 family, the proband has gait ataxia, titubation, ocular telangiectasias, elevated alpha-fetoprotein, absent IgA, low IgG2, and mild T cell lymphopenia. Clinical exome sequencing identified a *de novo* missense variant of uncertain significance (VUS) and a maternally inherited partial gene deletion in the *ATM* gene, but phase information for these variants could not be determined. Ataxia-telangiectasia is a recessive condition caused by *ATM* pathogenic variants, requiring two variants to be in *trans* to cause disease. Constellation, together with our pipeline, independently identified the *de novo* SNV, 11:108229319:C:G, and the precise breakpoints of the SV, NC_000011.11:g.108306295_108362948del. Furthermore, we were able to phase each variant on different haplotypes across the *ATM* gene, which enabled the identification of the *de novo* SNV occurring on the paternal haplotype, while the SV that is ∼77 Kb away, was shown to be maternally inherited (**Figure 5C, D**). Thus, Constellation identified the *ATM* SNV as likely pathogenic per ACMG/AMP variant curation guidelines for germline *ATM* sequence variants^31^, supporting the diagnosis of ataxia-telangiectasia, a neurodegenerative disorder with multiple emerging therapies^32,33^.

These findings underscore Constellation’s capacity to resolve clinically intractable cases by enhancing rare variant detection and phasing in clinically relevant contexts.

## Discussion

Rare disease genomics continues to face a critical gap: while genome sequencing has revealed thousands of pathogenic SNVs and small indels, many diagnoses remain unresolved because standard short-read DNA sequencing (SRS) misses structural variants (SV) and phasing^4^. Long-read sequencing addresses these challenges but remains limited by cost, DNA requirements, and thus scalability^9^. Here, we show that Illumina’s Constellation technology bridges this divide, delivering long-range information within standard short-read workflows and directly resolving or solving previously unsolved rare disease trios. In our study, Constellation confirmed 9 solved cases and we highlighted two previously unsolved trios, uncovering causal alleles despite their complexity or adjacency to segmental duplications. These included compound heterozygous events, phased de novo mutations, and SV within medically relevant genes.

While long reads remain necessary for large tandem repeat expansions^6^, ascertaining breakpoints in ultra-long SDs tandem arrays, and direct methylation profiling^8^, we demonstrate that Constellation captures the majority of phasing and SV events that underlie diagnostic uncertainty. This is achieved through leveraging proximity information between read clusters on the flow cell, which improves resolution of variants in repetitive regions and determines the allelic phase of variants, which are all central to rare disease diagnosis. Most impressively, this was shown over the *MECP2* complex rearrangements that could have been resolved with visual inspection based on Constellation data.

The overall result is a significant expansion of what SRS can deliver. Unlike previous long-range approaches, Constellation requires no special preparations and operates directly on existing Illumina infrastructure. This makes the method inherently scalable and easily transferable, enabling rapid adoption across sequencing centers and hospitals worldwide. By lowering technical and logistical barriers, Constellation extends long-range genomic insights to settings where long-read sequencing remains impractical, like due to the minimal amount of input DNA^3,9^. Its cost-efficiency supports routine use translational rare disease research, carrier screening, and population cohorts, where phasing and structural variants are crucial for interpretation but underrepresented with SRS. Additionally, its ability to derive long-range haplotype information directly from SRS data enables resolution of compound heterozygous variants in cases where parental samples are not available. As such, we show compelling cases that Constellation is a significant step to close the gap compared to long read sequencing and illuminates a larger part of the dark genome^4^ (e.g., *STRC/PMS2* coverage; *MECP2* junctions and CNV alterations) and its hidden variants in rare and potentially other diseases at scale.

The most striking findings in this study were the detection of *MECP2* complex rearrangements with multiple breakpoint junctions *in cis*, some of which map to segmental duplications, as well as the case that highlighted an SNV and SV in the compound heterozygous relationships. These cases obviously highlight the ability of Constellation, but more importantly, highlight the necessity to incorporate all variant types in genetic analysis. Obviously, there can be multiple reasons for failing to identify a causative variant in a rare disease case^34^. Some are related to candidate disease genes, mechanisms, or regions of the genome not represented in the reference genome^3,34,35^. Nevertheless, we hypothesize that a good proportion is because current diagnostic pipelines are weighted heavily toward SNVs and indels, leaving SV and phasing information underrepresented. This is also exemplified by lack of annotation or population frequency databases for SV^36^. Our results emphasize that comprehensive diagnosis of rare diseases and potentially other diseases requires equal consideration of SNVs, SVs, repeat expansions, and their combined effects. Clearly, all these variants coexist in our genomes and thus necessitate the simultaneous ascertainment of their impact. Constellation is, as such, another significant step towards this future, together with pangenomic analysis support such as DRAGEN.

In summary, Constellation transforms short-read sequencing into a platform capable of resolving long-range variant classes critical for diagnosis. By improving phasing, SV detection, and compound heterozygote variant resolution at scale, it significantly reduces the diagnostic gap and provides a cost-efficient path to wider adoption of precision genomics.

## Methods

### Consent agreements

All individuals or their guardians provided written informed consent for genomic studies and publication of clinical history under Baylor College of Medicine Institutional Review Board approved protocol H-29697.

### Constellation sequencing and data processing

#### Sample preparation and sequencing

**Sample preparation:** The DNA samples were quantified using the Qubit dsDNA BR Assay Kit (Thermo Fisher Scientific Q32853), and the DNA size was determined using automated pulsed-field capillary electrophoresis on the Agilent Femto Pulse instrument. Samples with Genomic Quality Number (GQN) > 5.0 (threshold of 10 kb) were considered suitable for Constellation sequencing. The GQN measures the integrity of DNA on a scale of 1 to 10 based on a threshold size. At a threshold of 10 kb, a GQN > 5.0 implies that at least 50% of the DNA fragments are >10 kb in size. The samples were diluted to 10 ng/μl or 30 ng/μl.

**Sequencing:** WGS using Constellation technology (For Research Use Only) was performed on GIAB control samples(4 HG002 replicates and the corresponding father HG003 and mother HG004) and all rare disease trios. Each genomic DNA sample (350 ng) was processed for sequencing according to the manufacturer’s protocol for the Illumina Constellation assay (Illumina, San Diego, CA). Briefly, each sample DNA (350 ng) mixture was added to a single well on the 8 sample library strip tube and loaded into the sequencing cartridge for the subsequent on-board tagmentation reaction. In addition to the genomic DNA, two additional reagents (FP1, FT2) were added to the CP1 and CP2 empty reservoirs of the NovaSeq X sequencing cartridge as part of the Constellation workflow. Sequencing was carried out on a NovaSeq X Plus system (Illumina) using NovaSeq X 10B flow cells (NovaSeq X Series 10B Reagent Kit, 300 cycles) with a custom Constellation recipe for use with NovaSeq X Series Control Software 1.3.0 for 150 bp paired-end reads.

#### Variant calling

All Constellation samples were demultiplexed by DRAGEN (v3.7.8) with option *––bcl-conversion-only true*. Samples were then uploaded to ICA for analysis with the TRIP-APP-KOL-Constellation_0-3_rc6 pipeline with default settings. This pipeline is an in-development version of the final Constellation pipeline that represents an early state of the final secondary analysis software package. In brief, the pipeline leverages proximity information to link nearby reads on the flow cell surface during mapping using DRAGEN^21^. A genome-wide colocation matrix is then generated from the aligned reads. Small variants are called from the aligned reads using the standard DRAGEN germline small variant caller. Variant and read phasing are then performed using HAPCUT2^38^ with the small variant calls and BAMs as input. The phased reads are then provided to DRAGEN for SV and CNV calling, which leverage proximity information to enhance variant calling accuracy. The VCF files of SNV/indels and SV calling results were directly downloaded from ICA for benchmarking.

#### Variant annotation and prioritization

**SNV and SV annotation:** The Constellation variant calling results were annotated in our in-house script for downstream filtering and pathogenic variants screening. SNVs and indels were annotated using ANNOVAR^39^ (v3.5) with multiple GRCh38-based reference databases, including refGene^40^, refGeneWithVer, avsnp151^41^, ClinVar^42^ (release 20240611), gnomAD v4.1 genome^43^, gnomAD v2.1.1 exome, dbNSFP^44^ v4.7a, dbscSNV^45^ v1.1, and cytoBand. SVs were annotated using AnnotSV^46^ v3.1.1 with the GRCh38 reference genome, incorporating RefSeq transcripts, cytobands, regulatory elements, and comprehensive pathogenicity resources, including ClinVarr^42^, ClinGen^47^, dbVar, DGV^48^, and 1000 Genomes^49^, together with intolerance and dosage-sensitivity metrics from ACMG^50^ (2023), ClinGen (2024), ExAC^51^, gnomAD, GenCC^52^ (2024), and OMIM^53^ (2024), as well as repeat content, GC composition, segmental duplications, and ENCODE^54^ blacklist regions.

**General filtering and pathogenic variants screening:** Compound heterozygous variant candidates were identified from annotated VCF files using a custom Python script that parsed the ANNOVAR^39^ trio annotation output TSV file. Variants were first filtered to retain only rare alleles (gnomAD v4.1^43^ genome allele frequency <0.001 or absent). Genotypes were normalized, and inheritance was evaluated under the compound heterozygosity model, requiring the proband to be heterozygous at each site with one allele transmitted from the mother and the other from the father, while excluding sex chromosome variants. Genes harboring at least one qualifying variant on each parental haplotype were reported as compound heterozygous candidates, with the contributing variants output for downstream interpretation.

De novo and Mendelian-error (MER) variant candidates were found using a custom Python script from ANNOVAR^39^ output, where the population allele frequency requirement and genotype normalization steps are still applied. Variant candidates should overlap with the exons of genes or be labeled as non-synonymous by ClinVarr^42^. Variants were retained only if the proband carried at least one alternate allele. De novo candidates were defined as heterozygous or homozygous alternate genotypes in the proband with both parents homozygous reference, whereas Mendelian error variants were flagged when observed genotypes violated Mendel’s laws of inheritance.

**Unsolved trio pathogenic variants prioritization and interpretation:** For unsolved cases, each trio has its specific clinical phenotype and an associated gene prioritization list (**Supplementary Table S4**). Therefore, we applied each personalized filtering script to each family. One universal rule for potential pathogenic variants is that the population allele frequency is <0.001, and the proband had to be genotyped. For the BCM17688_1 sample, a de novo missense VUS in the *ATM* gene was expected. Then we applied the previous script (see code availability) for de novo and MER variant screening with a specific requirement on overlapping with the *ATM* gene. For the BH8977_1 sample, it shared the same phenotype as BH8977_4 (sibling), in which case we should look for a homozygous variant overlapping with the clinician-provided gene list and shared by both of them. Then we required proband and BH8977_4 had to be genotyped, and parents cannot be homozygous alternative allele (ALT). Besides, as AD (Autosomal Dominant) and AR (Autosomal Recessive) genes require different numbers of variants (AD≥1, AR≥2) to be causative genes, we add a count match requirement for each gene. For the BCM17687 family, we expected either compound heterozygous variants or a de novo variant on the same gene list as the BH8977 family, in which case we applied the previous two scripts with specific requirements on gene match and variant count match. For the BH13518 family, the clinician suggested possible homozygous or compound heterozygous variants in *ETFA, ETFB, ETFDH, FLAD1, SLC52A1, SLC52A2, or SLC52A3* genes. Hence, we applied the previous two scripts (see code availability) with a specific gene overlap requirement. For the BCM17304 family, the pathogenic variant should be located on the *DMD* gene, where we applied scripts to screen candidates.

***MECP2* analysis:** From the colocation matrix of each sample, we manually checked the breakpoints in the *MECP2* region and checked all possible combinations of these breakpoints based on the colocation information brought by Constellation. Then, all possible scenarios were compared to the previously reported genome structure of each affected proband to assess the performanceability of Constellation on complex variants.

#### Standard Illumina short-read

**Variant calling:** The BAM file was downsampled and realigned to the GRCh38 reference with 65x coverage to maintain consistency with the observed coverage of Constellation samples. BWA mem^55^ (v0.7.17) was used to align short-read data with the *-M* option to mark shorter split hits as secondary. Two separate analysis strategies were applied for variant detection. In the first strategy, the BAM file was uploaded to ICA and went through the DRAGEN_Germline_Whole_Genome_4-4-4 analysis pipeline with *––enable-map-align, ––enable-duplicate-marking, ––enable-variant-caller, ––vc-enable-vcf-output, ––enable-sv, ––repeat-genotype-enable,* and *––enable-variant-annotation* options set as True. All variant calling results were then directly downloaded from ICA. In the second strategy, the aligned BAM file was run on Manta^24^ v1.6.0 germline mode with default settings for SV calling, and was run on DeepVariant^20^ v1.9.0 with *––model_type WGS* option for SNV/indel calling.

#### Oxford Nanopore

**Data collection:** the ONT data was downloaded from GIAB (see data availability), where two runs were merged using SAMtools (version 1.21) merge. Then we realigned it to the GRCh38 reference at 25x coverage.

**Variant calling:** The ONT R10 were also downsampled and realigned to the GRCh38 reference at 25x coverage. Minimap2^56^ (v2.28) was used to align long-read data with *-Y*, *-H*, *-a*, and *––MD* options, as well as technology-specific parameters (*-x map-hifi* and *-x map-ont*). Resulting alignments were converted to BAM format, sorted, and indexed using SAMtools^55^ (v1.19.2). Sniffles2(v2.4) with default setting was used to detect SV^5^ and Clair3^22^ v1.0.10 was used to detect small variants with technology-specific parameters (*-platform=“ont”, ––model_path=r1041_e82_400bps_sup_v500*) to call SNV/indels.

#### PacBio

**Data collection:** the PacBio dataset was downloaded from GIAB(see data availability)

**Variant calling:** The PacBio HiFi BAM file, was directly downloaded from GIAB (see Data availability section), where we downsampled it and realigned it to the GRCh38 reference at 25x coverage.

Sniffles2^5^(v2.4) with default setting was used to detect SV and Clair3^22^ v1.0.10 was used to detect small variants with technology-specific parameters ( *––platform=“hifi”, ––model_path=hifi_revio*) to call SNV/indels.

#### Variant phasing

We use WhatsHap^57^ (v2.8) to phase variants for the sequencing technologies except for Constellation technology, as its pipeline already reports phasing information in its VCF. We run the WhatsHap phase command with ––ignore-read-groups option. The VCF and corresponding BED files of benchmark sets were directly downloaded from the GIAB FTP site. For the SNV/indels benchmark, GIAB v4.2.1, T2T-Q100 v1.1, and CMRG v1.0 were considered. For the SV benchmark, T2T-Q100 v1.1 and CMRG v1.0 were considered.

#### HG002-based benchmarking

For SNV/indels benchmarking, we stratified the evaluation into four categories: SNV-only with genotype requirement (wtGT), SNV-only without genotype requirement (woGT), SNV+indels wtGT, and SNV+indels woGT. We used bcftools^58^ v1.19 with the –v snps option on SNV/indel calling VCF files to only select SNVs. When requiring genotype match, we run rtg vcfeval ––ref-overlap –b $base_vcf ––bed-regions ${reference_specific_bed} –c $comp_vcf ––vcf-score-field QUAL to evaluate the called SNV/indels by each caller against each reference set using rtgtools^59^(v3.13). When not requiring genotype match, we add ––squash-ploidy parameter to omit the GT requirement.

For SV benchmarking, we executed truvari bench *––refdist 2000 ––pctseq 0.7 ––pctsize 0.7 ––pctovl 0.0 ––passonly ––sizemin 50 ––sizefilt 50 ––sizemax 50000 ––pick ac ––extend 0 ––chunksize 5000 ––includebed ${reference_specific_bed} –b $ref_vcf –c $input_vcf*, followed by truvari refine *–reference ${reference.fa} ––regions ${candidate.refine.bed} ––use-original-vcfs ––coords R ––align mafft* to rigorously evaluate called SVs by each caller against each reference benchmark set in Truvari^60^ v5.3.1.

For phasing benchmarking, we collected metrics from WhatsHap stats (with the –block-list parameter) on all sequencing technologies on autosomes. Notably, to keep consistency of all sequencing technologies in the benchmark, we also use the WhatsHap metrics to summarize the Constellation phasing information. The percentage of phased SNV was directly reported from WhatsHap stats. The mean value of NG50 and the percentage of fully phased genes and SNVs by a single block were calculated by in-house scripts from WhatsHap output, which can be found in the Data Availability section. The percentage of genes that were phased by single phase block was calculated using BEDTools^61^ v2.30.0 with *-f 1* option to require full overlap between phase block reported by WhatsHap stats and our gene list (see Code availability).

### Data availability

The data used in this study are listed in the **Supplementary Table S5.**

GIAB benchmark sets:

Genome-wide:

https://ftp-trace.ncbi.nlm.nih.gov/ReferenceSamples/giab/data/AshkenazimTrio/analysis/NIST_HG002_DraftBenchmark_defrabbV0.019-20241113/

https://ftp-trace.ncbi.nlm.nih.gov/ReferenceSamples/giab/release/AshkenazimTrio/HG002_NA24385_son/NISTv4.2.1/GRCh38/

Medical regions:

https://ftp-trace.ncbi.nlm.nih.gov/ReferenceSamples/giab/release/AshkenazimTrio/HG002_NA24385_son/CMRG_v1.00/

PacBio WGS HG002:

https://ftp.ncbi.nlm.nih.gov/ReferenceSamples/giab/data/AshkenazimTrio/HG002_NA24385_son/PacBio_HiFi-Revio_20231031/HG002_PacBio-HiFi-Revio_20231031_48x_GRCh38-GIABv3.bam

ONT WGS HG002: https://epi2me.nanoporetech.com/giab-2025.01/

Illumina Standard Short-read Sequencing WGS HG002: https://precision.fda.gov/challenges/10/intro

All VCF files used in HG002 benchmark were uploaded to Zenodo: https://zenodo.org/records/17573936

### Code availability

Scripts for the analysis are available at https://github.com/jamesc99/Constellation

## Acknowledgments

FJS, SC, and XZ are partially supported by NIH grants (1U01HG011758-01, 1UG3NS132105 and R01HG011774)

## Contributions

**Sample collection and processing:** S.N.J. and D.G.C. coordinated sample curation and sequencing. Y.H., D.M.M., H.H.M., H.H.D., and K.P.B. oversaw sample preparation and sequencing. J.C.W. reviewed sample quality and sequencing metrics. D.G.C. and C.M.B.C. contributed to sample enrollment and interpretation.

**Data generation and analysis:** S.C. and Q.Z. performed data analysis and contributed to writing. Z.M.K. and J.R.F. contributed to data generation and computational analysis. X.Z. contributed to data interpretation and writing.

**Conceptualization, supervision, and writing:** F.J.S., D.G.C., and J.H. conceived and supervised the study, led project planning, and contributed to writing the manuscript. M.A.B. and A.C. contributed to study design and planning. J.E.P. and R.A.G. contributed to manuscript editing.

All authors reviewed and approved the final manuscript.

## References

1. Goodwin, S., McPherson, J. D. & McCombie, W. R. Coming of age: ten years of next-generation sequencing technologies. Nat Rev Genet 17, 333–351 (2016).

2. Pandey, R. et al. A meta-analysis of diagnostic yield and clinical utility of genome and exome sequencing in pediatric rare and undiagnosed genetic diseases. Genet Med 27, 101398 (2025).

3. Wojcik, M. H. et al. Beyond the exome: What’s next in diagnostic testing for Mendelian conditions. Am J Hum Genet 110, 1229–1248 (2023).

4. Sedlazeck, F. J., Lee, H., Darby, C. A. & Schatz, M. C. Piercing the dark matter: bioinformatics of long-range sequencing and mapping. Nat Rev Genet 19, 329–346 (2018).

5. Smolka, M. et al. Detection of mosaic and population-level structural variants with Sniffles2. Nat Biotechnol 42, 1571–1580 (2024).

6. English, A. C. et al. Analysis and benchmarking of small and large genomic variants across tandem repeats. Nat Biotechnol 43, 431–442 (2025).

7. Mahmoud, M., Agustinho, D. P. & Sedlazeck, F. J. A Hitchhiker’s Guide to long-read genomic analysis. Genome Res 35, 545–558 (2025).

8. Fu, Y., Timp, W. & Sedlazeck, F. J. Computational analysis of DNA methylation from long-read sequencing. Nat Rev Genet 26, 620–634 (2025).

9. Coster, W. D., De Coster, W., Weissensteiner, M. H. & Sedlazeck, F. J. Towards population-scale long-read sequencing. Nature Reviews Genetics vol. 22 572–587 Preprint at 10.1038/s41576-021-00367-3 (2021).

10. Arslan, S. et al. Sequencing by avidity enables high accuracy with low reagent consumption. Nat Biotechnol 42, 132–138 (2024).

11. Simmons, S. K. et al. Mostly natural sequencing-by-synthesis for scRNA-seq using Ultima sequencing. Nat Biotechnol 41, 204–211 (2023).

12. Chen, Z. et al. Ultralow-input single-tube linked-read library method enables short-read second-generation sequencing systems to routinely generate highly accurate and economical long-range sequencing information. Genome Res 30, 898–909 (2020).

13. Majidian, S., Agustinho, D. P., Chin, C.-S., Sedlazeck, F. J. & Mahmoud, M. Genomic variant benchmark: if you cannot measure it, you cannot improve it. Genome Biol 24, 221 (2023).

14. Dawood, M., et al. GREGoR: Accelerating Genomics for Rare Diseases. ArXiv (2024).

15. Chemistry Technical Document. Oxford Nanopore Technologies https://nanoporetech.com/document/chemistry-technical-document (2017).

16. PacBio library instructions. https://www.pacb.com/wp-content/uploads/Procedure-Checklist-Preparing-HiFi-SMRTbell-Libraries-using-SMRTbell-Express-Template-Prep-Kit-2.0.pdf.

17. Wagner, J. et al. Benchmarking challenging small variants with linked and long reads. Cell Genom 2, (2022).

18. Hansen, N. F., et al. A complete diploid human genome benchmark for personalized genomics. bioRxiv (2025) doi:10.1101/2025.09.21.677443.

19. Wagner, J. et al. Curated variation benchmarks for challenging medically relevant autosomal genes. Nat Biotechnol 40, 672–680 (2022).

20. Poplin, R. et al. A universal SNP and small-indel variant caller using deep neural networks. Nat Biotechnol 36, 983–987 (2018).

21. Behera, S. et al. Comprehensive genome analysis and variant detection at scale using DRAGEN. Nat Biotechnol 43, 1177–1191 (2025).

22. Zheng, Z. et al. Symphonizing pileup and full-alignment for deep learning-based long-read variant calling. Nat Comput Sci 2, 797–803 (2022).

23. Mahmoud, M. et al. Structural variant calling: the long and the short of it. Genome Biol 20, 246 (2019).

24. Chen, X., et al. Manta: rapid detection of structural variants and indels for germline and cancer sequencing applications. Bioinformatics 32, 1220–1222 (2016).

25. VCV000982662.10 – ClinVar – NCBI. https://www.ncbi.nlm.nih.gov/clinvar/variation/982662/.

26. Grochowski, C. M. et al. Inverted triplications formed by iterative template switches generate structural variant diversity at genomic disorder loci. Cell Genom 4, 100590 (2024).

27. Pehlivan, D. et al. Comprehensive assessment reveals numerous clinical and neurophysiological differences between MECP2-allelic disorders. Ann. Clin. Transl. Neurol. 12, 433–447 (2025).

28. Martin, A. R. et al. PanelApp crowdsources expert knowledge to establish consensus diagnostic gene panels. Nat Genet 51, 1560–1565 (2019).

29. Walker, L. C. et al. Using the ACMG/AMP framework to capture evidence related to predicted and observed impact on splicing: Recommendations from the ClinGen SVI Splicing Subgroup. Am. J. Hum. Genet. 110, 1046–1067 (2023).

30. Jaganathan, K. et al. Predicting Splicing from Primary Sequence with Deep Learning. Cell 176, 535–548.e24 (2019).

31. Richardson, M. E. et al. Specifications of the ACMG/AMP variant curation guidelines for the analysis of germline ATM sequence variants. Am J Hum Genet 111, 2411–2426 (2024).

32. Lynch, M. et al. Phase 2a/b randomised placebo-controlled dose-escalation trial of triheptanoin for ataxia-telangiectasia: treating mitochondrial dysfunction with anaplerosis. EBioMedicine 118, 105840 (2025).

33. Kim, J. et al. A framework for individualized splice-switching oligonucleotide therapy. Nature 619, 828–836 (2023).

34. Sullivan, J. A., Schoch, K., Spillmann, R. C. & Shashi, V. Exome/Genome Sequencing in Undiagnosed Syndromes. Annu Rev Med 74, 489–502 (2023).

35. Marwaha, S., Knowles, J. W. & Ashley, E. A. A guide for the diagnosis of rare and undiagnosed disease: beyond the exome. Genome Med 14, 23 (2022).

36. Zheng, X., et al. STIX: Long-reads based accurate structural variation annotation at population scale. bioRxiv (2024) doi:10.1101/2024.09.30.615931.

37. Chen, S. Ultrafast one-pass FASTQ data preprocessing, quality control, and deduplication using fastp. Imeta 2, e107 (2023).

38. Bansal, V. HapCUT2: A Method for Phasing Genomes Using Experimental Sequence Data. Methods Mol Biol 2590, 139–147 (2023).

39. Wang, K., Li, M. & Hakonarson, H. ANNOVAR: functional annotation of genetic variants from high-throughput sequencing data. Nucleic Acids Res. 38, e164 (2010).

40. Goldfarb, T. et al. NCBI RefSeq: reference sequence standards through 25 years of curation and annotation. Nucleic Acids Res. 53, D243–D257 (2025).

41. Sherry, S. T. et al. dbSNP: the NCBI database of genetic variation. Nucleic Acids Res. 29, 308–311 (2001).

42. Landrum, M. J. et al. ClinVar: improving access to variant interpretations and supporting evidence. Nucleic Acids Res. 46, D1062–D1067 (2018).

43. Atkinson, E. G. et al. Discordant calls across genotype discovery approaches elucidate variants with systematic errors. Genome Res. 33, 999–1005 (2023).

44. Liu, X., Li, C., Mou, C., Dong, Y. & Tu, Y. dbNSFP v4: a comprehensive database of transcript-specific functional predictions and annotations for human nonsynonymous and splice-site SNVs. Genome Med. 12, 103 (2020).

45. Jian, X. & Liu, X. In silico prediction of deleteriousness for nonsynonymous and splice-altering single nucleotide variants in the human genome. Methods Mol. Biol. 1498, 191–197 (2017).

46. Geoffroy, V. et al. AnnotSV: an integrated tool for structural variations annotation. Bioinformatics 34, 3572–3574 (2018).

47. Rehm, H. L. et al. ClinGen––the clinical genome resource. N. Engl. J. Med. 372, 2235–2242 (2015).

48. MacDonald, J. R., Ziman, R., Yuen, R. K. C., Feuk, L. & Scherer, S. W. The Database of Genomic Variants: a curated collection of structural variation in the human genome. Nucleic Acids Res. 42, D986–92 (2014).

49. 1000 Genomes Project Consortium et al. A global reference for human genetic variation. Nature 526, 68–74 (2015).

50. Richards, S. et al. Standards and guidelines for the interpretation of sequence variants: a joint consensus recommendation of the American College of Medical Genetics and Genomics and the Association for Molecular Pathology. Genet. Med. 17, 405–424 (2015).

51. Lek, M. et al. Analysis of protein-coding genetic variation in 60,706 humans. Nature 536, 285–291 (2016).

52. DiStefano, M. T. et al. The Gene Curation Coalition: A global effort to harmonize gene-disease evidence resources. Genet. Med. 24, 1732–1742 (2022).

53. Hamosh, A., Scott, A. F., Amberger, J. S., Bocchini, C. A. & McKusick, V. A. Online Mendelian Inheritance in Man (OMIM), a knowledgebase of human genes and genetic disorders. Nucleic Acids Res. 33, D514–7 (2005).

54. ENCODE Project Consortium. An integrated encyclopedia of DNA elements in the human genome. Nature 489, 57–74 (2012).

55. Li, H. & Durbin, R. Fast and accurate short read alignment with Burrows-Wheeler transform. Bioinformatics 25, 1754–1760 (2009).

56. Li, H. Minimap2: pairwise alignment for nucleotide sequences. Bioinformatics 34, 3094–3100 (2018).

57. Martin, M., et al. WhatsHap: fast and accurate read-based phasing. bioRxiv (2016) doi:10.1101/085050.

58. Danecek, P. et al. Twelve years of SAMtools and BCFtools. Gigascience 10, giab008 (2021).

59. Cleary, J. G., et al. Comparing Variant Call Files for performance benchmarking of next-generation sequencing variant calling pipelines. bioRxiv (2015) doi:10.1101/023754.

60. English, A. C., Menon, V. K., Gibbs, R., Metcalf, G. A. & Sedlazeck, F. J. Truvari: Refined structural variant comparison preserves Allelic diversity. bioRxiv (2022) doi:10.1101/2022.02.21.481353.

61. Quinlan, A. R. & Hall, I. M. BEDTools: a flexible suite of utilities for comparing genomic features. Bioinformatics 26, 841–842 (2010).

